# Which early indicator allows for a better understanding of the evolution of the COVID-19 epidemic in France?

**DOI:** 10.1101/2020.09.29.20203760

**Authors:** Patrice Loisel

## Abstract

We provide an explanation for the apparent discrepancy between the dynamic of the positive COVID-19 test rates and of the numbers of COVID-19-related hospital and intensive care admissions and deaths in France. We highlight the existence of a latency period of around 5 weeks between the infections of young individual (generally asymptomatic) to older individuals that may have a severe form of the disease and may be hospitalized. If the overall positive detection rate provided relevant information until the end of August, since the beginning of September the overall positive detection rate has reached a plateau and no longer provides relevant information on the current state of the epidemic. We show that the incidence rate in the 70+ age group is a relevant and early indicator of the epidemic. Furthermore, we have shown an identical doubling time of around 15 days for the following indicators: the incidence rate for the 70+ age group, the number of new admissions to hospital, intensive care unit admissions and deaths.

## 1 Introduction

The COVID-19 epidemic started in China in late 2019 in Hubei Province, then it spread around the world and became a pandemic [1]. A first wave reached France in late winter and continued in early spring 2020. After a lull at the end of spring following lockdown, since the end of July the epidemic has been restarting to form a second wave. This second wave is less strong than the first, in the sense that the evolution of the number of cases is less rapid. But we do not yet know the consequences in the medium term. In order to analyse this new phase of the epidemic we have data provided by Santé Publique France [2]. The weekly bulletin of Santé Publique France provides us with the the following indicators: new hospital admissions, new intensive care admissions and deaths.

From the beginning of the second wave, which can be estimated from the end of July to the beginning of September, these quantities provided us with relevant and consistent indicators to monitor the evolution of the epidemic: we could deduce a doubling of the quantities considered every 21 days. However, for the past month, there has been a discrepancy between the evolution of the rate of positive tests and the numbers of hospital admissions and deaths: while these numbers double every 16 days, in recent weeks the rate of positive tests has hardly changed at all. We will therefore try to explain this discrepancy. One possible explanation is the time lag between the infection dates of asymptomatic young people and the hospital admission dates of older people who develop severe forms, which might suggest that the epidemic is coming to an end. We will show that this is not exactly the case and that the positive tests rate is no longer a relevant indicator and will seek to propose a new indicator.

In section 2, based on the available indicators, we will put in the discrepancy between the positive tests rate evolution and the number of new hospital admissions. In section 3, we will use additional data based on age structure and positive tests and will propose a new indicator. To conclude, we will summarise the information we have been able to extract from the data.

## 2 Analysis of available indicators

Every week, Santé Publique France publishes a newsletter providing, among other things, the following indicators: positive tests rate, numbers of new hospital admissions, new intensive care admissions and deaths. We are interested in the second wave of the epidemic that began around end of July. We adjoin a complementary indicator: the incidence rate. In Table 1 we reproduce the data provided by Santé Publique France.

**Table 1:** Positive tests rate, incidence rate, numbers of hospital admissions, intensive care admissions and deaths

|  | *Week number* |  |  |  |  |  |  |  |  |  |
| --- | --- | --- | --- | --- | --- | --- | --- | --- | --- | --- |
|  | 29 | 30 | 31 | 32 | 33 | 34 | 35 | 36 | 37 | 38 |
| positive tests rate (in % ) | 1.23 | 1.42 | 1.58 | 2.17 | 3.14 | 3.81 | 4.44 | 5.42 | 5.45 | 5.79* |
| incidence rate (for 100000) | 6.63 | 9.77 | 12.6 | 17.6 | 25.7 | 41.7 | 59.0 | 78.2 | 95.5 | 110.2 |
| Nb of hospital admissions | 604 | 749 | 778 | 782 | 1008 | 1084 | 1337 | 1704 | 2464 | 3657 |
| Nb of intensive care admissions | 83 | 85 | 105 | 122 | 128 | 174 | 210 | 288 | 427 | 599 |
| Nb of deaths | 94 | 78 | 79 | 66 | 97 | 112 | 109 | 129 | 265 | 332 |

\*: data that may be actualized next week

The positive tests rate has been increasing exponentially from the end of July to the beginning of September, but in the last few weeks, this rate growth has slowed. To a lesser extent, we observe the same behaviour for the incidence rate. This could indicate a slowing down of the epidemic. To find out whether it is the case or not and to better understand the rate evolution, we make a preliminary evaluation of the local *k* parameter of the exponential describing the evolution of the indicators of interest and the corresponding doubling times. This will allow us to detect break points in the epidemic process. We give in Table 2 the growth rates over 3 weeks: 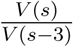 for the positive tests rate and the numbers of hospital admissions, intensive care admissions or deaths, we give an estimate of the associated doubling time:

**Table 2:** Local estimates of growth rate and doubling time for the positive tests rate, incidence rate, number of hospital admissions, number of intensive care admissions, number of deaths

|  |  |  |  |  |  |  |
| --- | --- | --- | --- | --- | --- | --- |
| *Positive tests rate* |  |  |  |  |  |  |
| Week ratio | 33/30 | 34/31 | 35/32 | 36/33 | 37/34 | 38/35 |
| rate of increase | 2.21 | 2.41 | 2.05 | 1.73 | 1.44 | 1.30* |
| local doubling time (in days) | **18.4** | **16.5** | **20.3** | **26.6** | **39.9** | **54.9\*** |
| *Incidence rate* |  |  |  |  |  |  |
| Week ratio | 33/30 | 34/31 | 35/32 | 36/33 | 37/34 | 38/35 |
| rate of increase | 2.63 | 3.32 | 3.343 | 3.05 | 2.29 | 1.87 |
| local doubling time (in days) | **15.1** | **12.1** | **12.1** | **13.1** | **17.6** | **23.3** |
| *Number of hospital admissions* |  |  |  |  |  |  |
| Week ratio | 33/30 | 34/31 | 35/32 | 36/33 | 37/34 | 38/35 |
| rate of increase | 1.35 | 1.39 | 1.71 | 1.69 | 2.27 | 2.74 |
| local doubling time (in days) | 49.0 | 43.9 | 27.1 | 27.7 | **17.7** | **14.5** |
| *Number of intensive care admissions* |  |  |  |  |  |  |
| Week ratio | 33/30 | 34/31 | 35/32 | 36/33 | 37/34 | 38/35 |
| rate of increase | 1.51 | 1.66 | 1.72 | 2.25 | 2.45 | 2.85 |
| local doubling time (in days) | 35.6 | 28.8 | 26.8 | 17.9 | **16.2** | **13.9** |
| *Number of deaths* |  |  |  |  |  |  |
| Week ratio | 33/30 | 34/31 | 35/32 | 36/33 | 37/34 | 38/35 |
| rate of increase | 1.24 | 1.42 | 1.65 | 1.33 | 2.37 | 3.05 |
| local doubling time (in days) | 66.8 | 41.7 | 29.0 | 51.1 | **16.9** | **13.1** |

Reading the results confirms for the positive tests rate and the incidence rate, a first phase between the end of July and the end of August. There is indeed an exponential growth with a doubling time of approximately 20 days followed by a period of stagnation for the positive tests rate and an exponential growth with a doubling time of approximately 15 days followed by a slowdown in growth. On the basis of the respective trends in doubling time for the positive test rate and incidence rate on the one hand and the number of hospital admissions on the other, it can be assumed (to be verified) that there is a latency period of the order of a few weeks between the rates and the number of hospital admissions. Furthermore, we note that :

- while the positive tests rate and incidence rate had an exponential growth the numbers of new admissions and deaths increased only marginally. This very weak evolution made some observers say that there was no cause for concern and that the increase in the rates was anecdotal because it only affected asymptomatic young people. This is not exactly the case because it means forgetting the existence of time lags, which we is described in the following.
- while in recent weeks the positive tests rate have stagnated and the growth of the incidence rate slowed down, the number of new admissions follows an exponential trajectory with a doubling time significantly lower than the rates in the previous phase: 15 days instead of 20 days. This is evidence that the epidemic is far from over.

To confirm this discrepancy between the doubling time of the positive test rate or the incidence rate and the current doubling time of new admissions we operate regressions of the log of positive tests rate, incidence rate, number of hospital admissions, intensive care admissions and deaths. We obtain the following results (Table 3) for the estimates of the *k* parameter of the exponential with the associated doubling time:

**Table 3:** Results of the regression of the log of positive tests rate, incidence rate, numbers of hospital admissions, intensive care admissions and deaths

| | $k$<br>(day<sup>-1</sup>) | Standard deviation<br>(day<sup>-1</sup>) | p-value | Doubling time<br>(day) |
| --- | --- | --- | --- | --- |
| positive tests rate (S29-36) | 0.033 | 0.002 | $1.77 \cdot 10^{-6}$ | 21.3 |
| Incidence rate (S29-36) | 0.051 | 0.001 | $2 \cdot 10^{-8}$ | 13.5 |
| Incidence rate (S34-38) | 0.035 | 0.003 | 0.002 | 20.0 |
| hospital admissions (S34-38) | 0.043 | 0.004 | 0.001 | **16.0** |
| intensive care admissions (S33-38) | 0.044 | 0.002 | $3.92 \cdot 10^{-5}$ | **15.8** |
| deaths (S34-38) | 0.044 | 0.010 | 0.024 | **15.8** |

The doubling time is around 21 days for the positive tests rate since week 29 and for the incidence rate since week 34 (after being around 15 days) while the doubling time for new hospital admissions and intensive care admissions is 16 days. The result concerning the growth rate for the number of deaths is not very significant, it will be necessary to wait another week to confirm the value obtained for the doubling time.

But then how can we explain the discrepancy in behaviour between the evolution of the positive tests rate or the incidence rate and the numbers of new admissions or deaths? To answer this question, we focus on the incidence rate and consider the evolution of the incidence rate by age group.

## 3 Analysis of the age structure of positive tests rates

Table 4 shows for each week of the incidence rate per 100 000 inhabitants for different age groups [3]. Figure 2 gives the contour plot of the incidence rate.

**Table 4:** Incidence rate per 100 000 for different age groups

| Age group | Week number |  |  |  |  |  |  |  |  |  |  |
| --- | --- | --- | --- | --- | --- | --- | --- | --- | --- | --- | --- |
|  | 28 | 29 | 30 | 31 | 32 | 33 | 34 | 35 | 36 | 37 | 38 |
| 0-9 | 3.48 | 3.36 | 4.26 | 4.66 | 6.40 | 7.66 | 10.7 | 16.0 | 24.8 | 38.6 | 34.9 |
| 10-19 | 5.36 | 5.76 | 8.67 | 11.6 | 15.1 | 23.1 | 37.2 | 56.4 | 82.5 | 109. | 136. |
| 20-29 | 9.78 | 11.0 | 19.6 | 30.8 | 45.5 | 70.3 | 127. | 177. | 213. | 231. | 259. |
| 30-39 | 8.17 | 9.99 | 14.1 | 18.5 | 26.9 | 37.4 | 61.3 | 86.2 | 110. | 122. | 139. |
| 40-49 | 6.90 | 7.53 | 10.5 | 11.8 | 17.5 | 26.3 | 40.6 | 55.8 | 76.7 | 98.3 | 114. |
| 50-59 | 5.61 | 6.45 | 8.75 | 10.9 | 13.6 | 21.2 | 32.7 | 45.9 | 63.2 | 84.5 | 97.0 |
| 60-69 | 4.37 | 4.77 | 6.82 | 7.75 | 9.77 | 13.3 | 21.1 | 29.1 | 41.9 | 52.3 | 63.3 |
| 70-79 | 3.63 | 4.72 | 6.62 | 6.09 | 9.04 | 11.5 | 13.7 | 21.0 | 28.9 | 40.5 | 49.2 |
| 80-89 | 3.54 | 3.88 | 5.45 | 6.47 | 9.18 | 10.8 | 13.3 | 19.2 | 30.3 | 44.7 | 56.2 |
| 90- | 6.57 | 6.25 | 8.99 | 9.64 | 13.7 | 15.6 | 19.6 | 29.0 | 47.1 | 86.2 | 101. |

From the Table 4 or the associated Figure 1, we notice that travelling waves start from the age group 20-29 years old: for example the incidence rate per 100 000 inhabitants (around 19.6) of the age group 20-29 years old in week 30 spreads to the age group 30-39 years old in week 31, … until the age group 70-79 years old in week 35. This phenomena is observed in the following weeks for higher rates. We deduce that the epidemic move from the age group 20-29 years old to the other class at a speed of 10 years per week until the age group 70-79 years old. Hence we have a first result:

(R1) : The latency period between the infections of young individual (generally asymptomatic) to older individuals that may have a severe form of the disease and may be hospitalized is close to 5 weeks.

**Figure 1:**
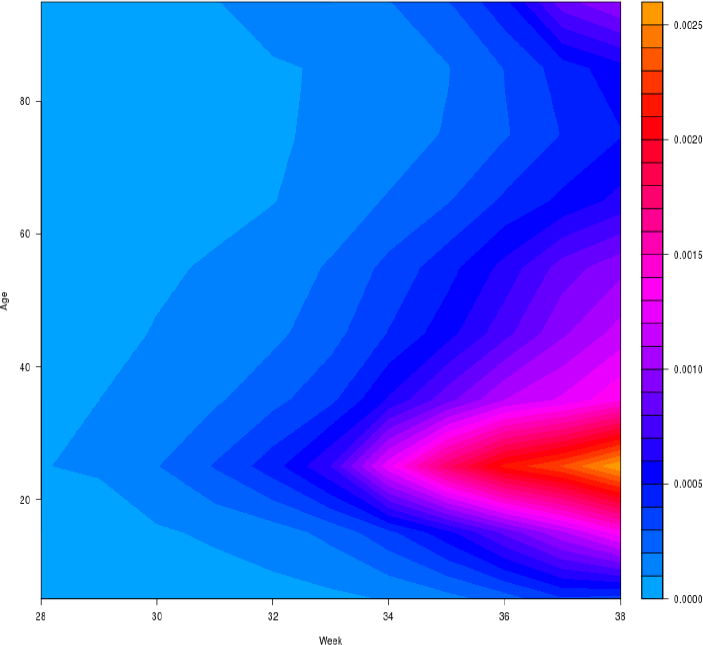
Contour plot of incidence rate for the different age groups

From :

- the vast majority of those who test positive are young people who will not develop serious forms of the disease.
- the test detects very recent infections
- hospitalisations mainly concern individuals of more than 70 years old

it can be deduced that the positive tests rate or the incidence rate at a given date will only lead to an increase in the number of new admissions 5 weeks later. This mini-conclusion is consistent with the facts we observed earlier: the exponential phase of the positive tests rate and incidence rate start in week 29 and that of new admissions only in week 33 or 34.

We still have to explain the second phase where there is a discrepancy: the positive detections rate stagnates and the incidence rate slowly increases while the numbers of new hospital admissions is growing exponentially at a higher rate. The answer lies in the process of the spread of the virus in the age groups. Indeed, the overall incidence rate (resp. positive tests rate) is an aggregation of incidence rates by age group and the structure of these rates in the age groups changes significantly over time. At the beginning of the second wave, it is mostly young people who have been contaminated and the more the weeks go by, the more older people are becoming infected. Since the end of August, the positive tests rate and incidence rate among older people have been increasing much faster than among young people: the incidence rate in the 70-79 (resp. 80-89) age group was multiplied by 3.6 (resp. 4.2) between weeks 34 and 38, whereas it was only multiplied by 2 for the 20-29 age group, by 2.6 for the total population. So even though the overall rate does not increase much anymore, the number of individuals who will develop severe forms is increasing very significantly. We then obtain our second and third results:

(R2): The relative discrepancy between overall incidence rate and number of hospital admissions is explained by the changing age structure of the population of individuals who test positive.

(R3): Thus if the overall positive tests rate provided relevant information until the end of August, since the beginning of September the overall positive tests rate no longer provides relevant information on the current state of the epidemic.

In order to know where we are in the evolution of the epidemic, it is therefore necessary to observe the incidence rate among the population at risk of developing a serious form, namely the age group of 70 years and over. To this end, we have calculated an incidence rate in this population, taking into account the numbers in the three age groups: 70-79, 80-89, 90 and over. We obtain the following evolution of the rates for the 70+ age group:

As before, we calculate local doubling times: :

We detect two phases, the first with a doubling time of around 20 days and the second with a doubling time of around 15 days. Once again we perform linear regressions and we get for the second phase:

Moreover in the last weeks, the evolution of the number of hospital admissions is quite similar to the evolution of the incidence rate for the 70+ age group.

Results of Table 7 thus confirms our forth and fifth results:

**Table 5:** Incidence rate for individuals older than 70 years

|  |  |  |  |  |  |  |  |  |  |  |
| --- | --- | --- | --- | --- | --- | --- | --- | --- | --- | --- |
| week number | 29 | 30 | 31 | 32 | 33 | 34 | 35 | 36 | 37 | 38 |
| incidence rate 70+ | 4.70 | 6.63 | 6.74 | 9.78 | 11.9 | 14.5 | 21.7 | 32.0 | 48.6 | 59.1 |

**Table 6:** Rate of increase of incidence rate and doubling time for individuals older than 70 years

|  |  |  |  |  |  |  |
| --- | --- | --- | --- | --- | --- | --- |
| week ratio | 33/30 | 34/31 | 35/32 | 36/33 | 37/34 | 38/35 |
| rate of increase | 1.79 | 2.15 | 2.22 | 2.70 | 3.36 | 2.73 |
| associated doubling time (in days) | 25.0 | 19.0 | 18.3 | **14.7** | **12.0** | **14.5** |

**Table 7:** Rate of increase of the incidence rate and doubling time for individuals older than 70 years

|  |  |  |  |  |
| --- | --- | --- | --- | --- |
| | $k$<br>(day<sup>-1</sup>) | Standard deviation<br>(day<sup>-1</sup>) | p-value | Doubling time<br>(day) |
| Incidence rate 70+ (S33-38) | 0.049 | 0.003 | 4.9 10<sup>-5</sup> | **14.1** |

(R4) : The incidence rate in the age group 70 and over is currently an early indicator of the epidemic.

(R5) : The doubling times of the incidence rate for the 70+ age group, the number of new admissions to hospital, intensive care unit admissions and deaths are around 15 days.

These results will have to be confirmed by the data of the following week.

## 4 Conclusion

We notice a discrepancy between the dynamic of the positive tests rate and the dynamics of the numbers of hospital admissions, intensive care admissions and deaths. These three quantities double every 15 days. The positive tests rate appears to have slowed grown over the past two weeks. We show that this apparent discrepancy is explained by the changing age structure of the population of individuals who test positive. We highlight the existence of a latency period of about 5 weeks between the positive detection rate and the number of new hospital admissions in the phase of the exponential dynamics of the positive tests rate. Thus if the overall positive tests rate provided relevant information until the end of August, since the beginning of September the evolution of the overall positive tests rate has reached a plateau and no longer provides relevant information on the current state of the epidemic. However, the incentive rate in the 70+ years old group is a relevant and early indicator of the epidemic. Furthermore, we have shown an identical doubling time of around 15 days for the following indicators: the incidence rate for the 70+ age group, the number of new admissions to hospital, intensive care unit admissions and deaths.

## Data Availability

All data is free of copyright

